# Personalised Cardiac Rehabilitation Outreach Service Reduces Mortality and Hospitalisations in Heart Failure with Reduced Ejection Fraction

**DOI:** 10.1101/2021.12.05.21267214

**Authors:** Zaidon Al-Falahi, Giuseppe Femia, Linda Gardiner, Jodie Ekholm, Kishore Kadappu, Rohan Rajaratnam

## Abstract

**Background:** Heart failure (HF) has become a major cause of morbidity and mortality worldwide. Despite significant improvements in the management of HF, the overall outcomes remain poor. In addition to pharmacotherapy and device therapy, non-pharmacological interventions are needed to mitigate the effects of this illness. The aim of this study was to evaluate the impact of the heart failure outreach program on the rate of mortality, HF hospitalisations and guideline directed medical therapy (GDMT) for HF in South Western Sydney Local Health District (SWSLHD).

**Methods:** In this observational, registry based study, adult patients diagnosed with Heart failure with reduced ejection fraction (HFrEF) within the South Western Sydney Local Health District (SWSLHD) and invited to participate in the heart failure outreach service between March 2011 and January 2016 were included in the study. The primary outcome was all-cause mortality. In addition, we examined the rate of optimal medical therapy, HF hospitalisations and the total lengths of stay.

**Results:** A total of 818 patients were included in the study; 470 (57.5%) patients were enrolled and 348 (42.5 %) not enrolled into the program. At the end of the follow up period (median 978 days, interquartile range (IQR) 720-1237), the primary outcome of mortality was observed significantly less in the enrolled group (122 (26%) vs. 133 (38.2%), p<0.001) independently of other variables. In addition, significantly fewer enrolled patients had >3 hospital admissions for HF (16.2% vs. 35.6%, p<0.001) and reduced median admission days (14.5 days [IQR 8-25] vs 22 [IQR 12-37], p <0.001). Patients enrolled into the program were much more likely to be on GDMT (76.6% vs 56.6%, p<0.001).

**Conclusions:** Enrolment in the heart failure outreach program was associated with a significant reduction in mortality as well as a reduction in the frequency and length of hospital admissions. In addition, the rate of GDMT was significantly higher in the enrolled group. With the high prevalence of heart failure, these programs should be considered in the routine management of patients with HFrEF.

## Introduction

Heart failure is a major cause of cardiovascular morbidity and mortality and has been described as a worldwide pandemic [1]. The one year mortality for patients with heart failure ranges from 17-45%, with the majority dying within 5 years [2]. In addition, heart failure is associated with reduced quality of life and a significant healthcare cost burden, making tailored interventions that reduce morbidity, mortality and hospitalisations vital to patients, families and healthcare systems [3, 4]. While the prevalence of heart failure is only set to increase, improvement in heart failure outcomes appear to have plateaued in the past two decades and survival remains worse than that of prostate, bowel and breast cancers [5-8].

Pharmacotherapy, in addition to device therapy in selected patients, have been the cornerstone of management for heart failure with reduced ejection fraction (HFrEF) with improvement in mortality and hospitalisations [9]. In addition, a number of non-pharmacologic interventions have emerged such as multimodal, multidisciplinary cardiac rehabilitation programs [10]. Although trials, observational studies and systematic reviews have shown consistent safety, improvement in exercise capacity and overall quality of life, they have not shown a significant and consistent reduction in mortality [11-14]. One possible explanation is the heterogeneity of these programmes and the short duration of both the intervention and the follow up period. There appears to be a trend for mortality benefit with longer periods of follow up [15, 16], hence it is important to evaluate the role of heart failure programmes on long term outcomes.

The aim of this study was to evaluate the impact of the heart failure outreach program on all-cause mortality, hospitalisations and adherence with guideline directed medical therapy (GDMT) in South Western Sydney Local Health District. Given the available evidence, we hypothesised that beyond the symptomatic and quality of life benefits of cardiac rehabilitation in chronic heart failure, mortality benefits were more likely to be evident with longer periods of follow up.

## Methods

### The Heart Failure Outreach Service Intervention

This is a multidisciplinary home-based outreach service led by a specialist heart failure nurse. Routinely, adult patients treated for an episode of heart failure at any of the hospitals within the SWSLHD get further screened by a heart failure nurse upon discharge. Patients with reduced left ventricular ejection fraction on echocardiography are eligible for the program, their clinical details are entered into the heart failure registry and receive a written invitation for home-based cardiac rehabilitation. Their clinical details are entered into Non-responders get a phone call follow up invitation. Home visits are performed on regular intervals and consist of physical examination, fluid status assessment and medications review. Education is consistently provided regarding medications compliance, side effects management and support. Dietary advice and education is also provided, especially regarding salt and fluid restriction, including dietician home visits. Physical exercises are prescribed and performed under supervision, tailored to individual patient needs.

### Study design and patients selection

This is an observational, multi-centre, registry based study. All adult patients identified by the SWSLHD heart failure outreach service with reduced left ventricular ejection fraction (LVEF <50%) on echocardiography and admitted to hospital with an episode of symptomatic heart failure were invited to participate between March 2011 and January 2016 and are included in the analysis.

Baseline information collected included age, sex, height and weight, enrolment status, vital signs on initial review, basic biochemical profile (creatinine, electrolytes, haemoglobin levels), echocardiographic estimation of LVEF and pulmonary artery systolic pressure (PASP), smoking status, medication history and presence or absence of ischemic cardiomyopathy and atrial fibrillation (AF). This information was populated prospectively into the heart failure registry and is obtained from the electronic health record as well as patient interviews. Mortality data was obtained from multiple sources including the electronic health records, follow up data, and from the primary care centres where patients are enrolled. The data for this study was entered as part of routine care and accessed and linked with the ethics approval “Multicultural Emergency Medicine Epidemiology (MEME) Study, HREC: LNR/17/LPOOL/432; project number HE17/234, approved 15^th^ Jan2018 and amended Nov 2020”

### Outcomes and Definitions

The primary outcome was all-cause mortality.

Hospital admissions were defined as any admission where management of heart failure exacerbation was included as a principal diagnosis in the electronic health record.

GDMT includes beta-blockers, angiotensin converting enzyme inhibitor (ACE-I), angiotensin receptor blocker (ARB) and spironolactone. Optimal medical therapy (OMT) was defined as a combination of any three of Beta blockers, ACE-I or ARB and spironolactone.

### Statistical Analysis

All analysis was performed using the Statistical Package for Social Sciences (SPSS), GraphPad software and Microsoft Excel. Categorical variables were presented in numbers and percentage (%) and continuous variables were presented as mean ± standard deviation or median/interquartile range (IQR). Data was visualised using histograms, box and Q-Q Plots, and normality was tested by Kolmogorov-Smirnov and Shapiro-Wilkins tests. Fisher’s exact tests were performed to compare categorical variables between groups, Student T tests for continuous variables and Mann-Whitney U tests to compare medians. A p value <0.05 was considered statistically significant. Logistic regression analysis was performed for statistically significant factors associated with mortality, and Odds Ratios (OR) were calculated with 95% Confidence Intervals (CI). Furthermore, a Kaplan-Meier survival analysis was constructed from the available heart failure diagnosis follow up dates.

## Results

### Baseline characteristics

A total of 818 patients were included in the study; 470 (57.5%) patients were enrolled into the HF outreach program. The median age in the enrolled group was slightly younger than the non-enrolled group (81 years [IQR 73-87] vs 83 years [IQR 74-89], p <0.001). In the enrolled group there were slightly less females (40.6% vs. 50.6%, p = 0.055), lower mean left ventricular systolic function (median 33 [IQR 23-41] vs 35 [IQR 25-45], p 0.029) and lower median pulmonary artery systolic pressure in the enrolled group (40 mmHg [IQR 32-51] vs 41 mmHg [IQR 35-52], p = 0.028). Furthermore, the enrolled group had a significantly higher prevalence of ischemic cardiomyopathy (69.6% vs. 49.4%, p <0.001). Body mass index was comparable across the two groups, and there was no statistically significant difference in the prevalence of atrial fibrillation and smoking.

### Primary Outcomes

The median follow up period was 978 days [IQR 720 - 1237]. Over this period, enrolment in the clinic was associated with a significant reduction in mortality (26.0% vs 38.2%, p < 0.001). Using logistic regression analysis, the strongest predictor of survival was enrolment status (OR 0.606, CI 0.436-0.841, p 0.003), followed by statin therapy (OR 0.682, CI 0.495-0.939, p 0.019), an effect driven mainly by mortality benefit in the subgroup with ischemic cardiomyopathy (p <0.001). Conversely, the strongest predictor of mortality was non-enrolment (OR 1.65, CI 1.189-2.29, p 0.003) followed by the presence of ischemic cardiomyopathy (OR 1.533, CI 1.089-2.156, p 0.014). In addition, the following factors made small but statistically significant contributions to mortality: Older age (OR 1.033, CI 1.017-1.049, p <0.001), higher creatinine levels (OR 1.004, CI 1.003-1.005, p <0.001) and higher estimated pulmonary artery systolic pressures (OR 1.014, CI 1.002-1.027, p .014).

In addition, female sex (p < 0.001), higher haemoglobin levels (p = 0.039), ACE inhibitor therapy (p = 0.049) and smoking status (p = 0.018) had an effect on mortality on group analysis, but were no longer statistically significant when corrected in a logistic regression model (Table 3).

**Table 1.**
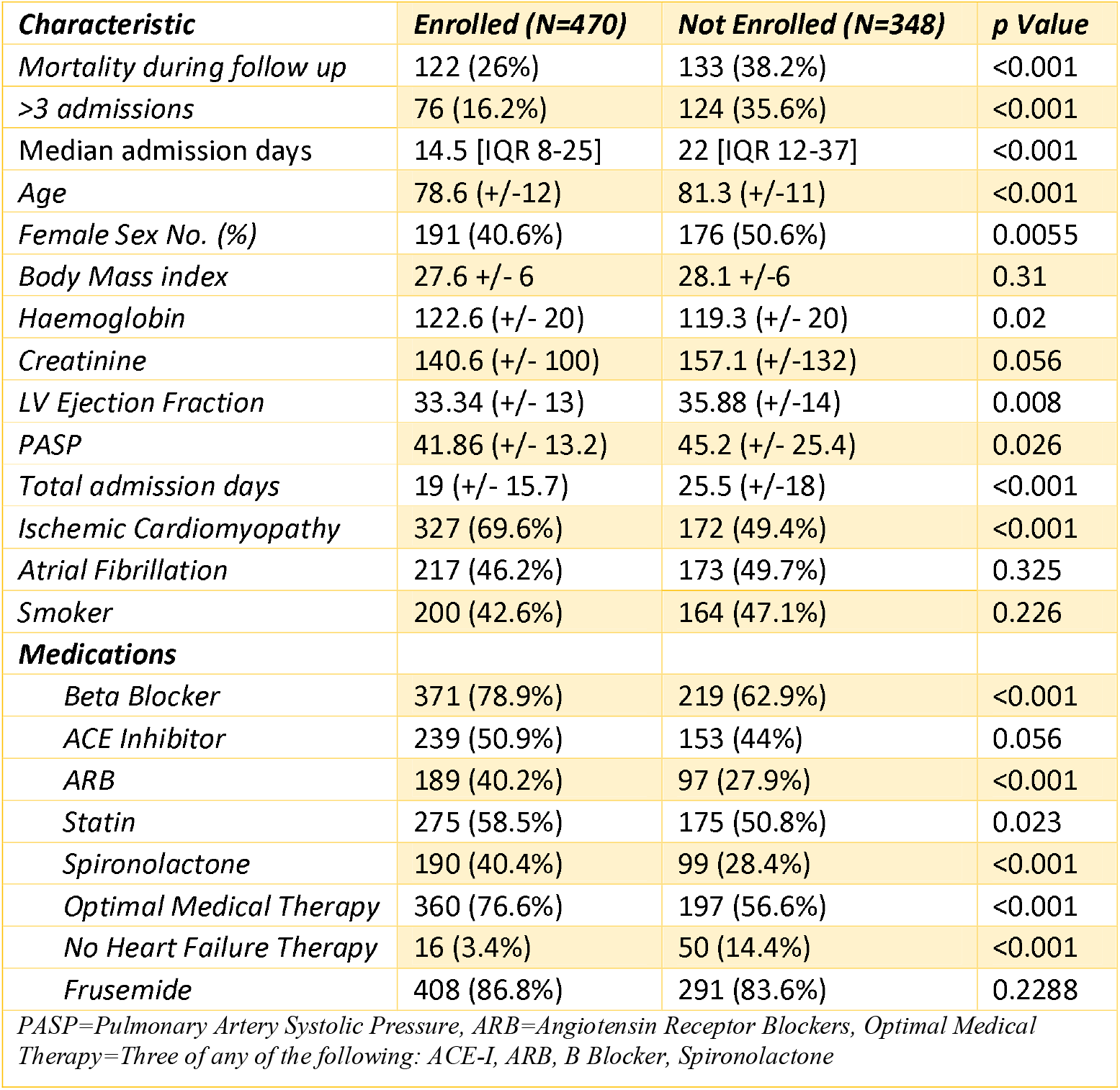
Characteristics by Enrolment.

**Table 2.** Characteristics By Mortality.

| **Characteristic** | **Deceased (N=255)** | **Alive (N=563)** | **p Value** |
| --- | --- | --- | --- |
| *Enrolment* | 122 (47.8%) | 348 (61.8%) | <0.001 |
| *Age* | 82.3 (+/- 10) | 78.6 (+/-12.1) | <0.001 |
| *Female Sex No. (%)* | 109 (42.7) | 258 (45.8) | <0.001 |
| *Body Mass Index* | 27.8 (+/- 6) | 27.9 (+/-6.6) | 0.969 |
| *Haemoglobin* | 119 (+/- 20) | 122.2 (+/-20.1) | 0.039 |
| *Creatinine* | 187 (+/-154) | 130 (+/-92) | <0.001 |
| *LV Ejection Fraction* | 34.1 (+/- 14.6) | 34.6 (+/-13) | 0.609 |
| *PASP* | 46.35 (+/- 29) | 42 (+/- 13) | 0.018 |
| *Total admission days* | 22.7 (+/- 17) | 21.3 (+/-17.2) | 0.289 |
| *Ischemic Cardiomyopathy* | 167 (65.5%) | 332 (59%) | 0.089 |
| *Atrial Fibrillation* | 123 (48.2%) | 267 (47.5%) | 0.88 |
| *Smoker* | 130 (50.1%) | 234 (41.2%) | 0.018 |
| **Medications** |  |  |  |
| *Beta Blocker* | 174 (68.2%) | 416 (73.9%) | 0.12 |
| *ACEI* | 109 (42.7%) | 283 (50.3%) | 0.049 |
| *ARB* | 89 (34.9%) | 197 (35%) | 1 |
| *Spironolactone* | 81 (31.8%) | 208 (36.9%) | 0.156 |
| *OMT* | 164 (64.3%) | 393 (69.8%) | 0.124 |
| *No Heart Failure Therapy* | 28 (11%) | 38 (67%) | 0.052 |
| *Frusemide* | 221 (86.7%) | 478 (84.9%) | 0.593 |
| *Aspirin* | 142 (55.7%) | 326 (57.9%) | 0.592 |
| *Digoxin* | 47 (18.4%) | 94 (16.7%) | 0.549 |
| *Statin* | 123 (48.2%) | 327 (58.1%) | <0.001 |
| *- ICM* | 84 | 204 | <0.001 |
| *- NICM* | 39 | 123 | 0.167 |
| *PASP=Pulmonary Artery Systolic Pressure, ARB=Angiotensin Receptor Blockers, OMT= Optimal Medical Therapy=Three of any of the following: ACE-I, ARB, B Blocker, Spironolactone, ICM= Ischemic cardiomyopathy, NICM=Non-ischemic cardiomyopathy* |  |  |  |

**Table 3.** Logistic Regression.

| **Mortality** | **Odds Ratio (95% C.I.)** | **P Value** |
| --- | --- | --- |
| **Enrolment in program** | 0.606 (0.436-0.841) | 0.003 |
| **Age** | 1.033(1.017-1.049) | <0.001 |
| **Female sex** | 0.765 (0.553-1.059) | 0.107 |
| **Ischemic Cardiomyopathy** | 1.533 (1.089-2.156) | 0.014 |
| **Smoking** | 1.330 (0.968-1.828) | 0.079 |
| **Haemoglobin levels** | 0.999 (0.991-1.007) | 0.795 |
| **Creatinine levels** | 1.004 (1.003-1.005) | <0.001 |
| **PASP** | 1.014 (1.002-1.027) | 0.023 |
| **ACE-I therapy** | 0.886 (0.644-1.219) | 0.456 |
| **Statin therapy** | 0.682 (0.495-0.939) | 0.019 |

### Other outcomes

Enrolment was also associated with significantly lower rate of recurrent HF admissions (>3 admissions 16.2% vs 35.6%, p <0.001) and reduced median admission days (14.5 days [IQR 8-25 ] vs 22 days [IQR 12-37], p <0.001).

Enrolment in the program was strongly associated with a rate of GDMT including beta-blockers (78.9 % vs 62.9%, p < 0.001), ACE-I (50.9% vs 44%, p 0.056), ARB (40.2% vs 27.9%, p < 0.001) and spironolactone (40.4% vs 28.4%, p < 0.001). The rate of optimal medical therapy was significantly more prevalent in the enrolled group (76.6% vs 56.6%, p <0.001); lack of GDMT was observed far less frequently in the enrolled group (3.4% vs 14.4%, p <0.001). Frusemide intake was comparable across both groups (86.8% in the enrolled groups vs 83.6% in the non-enrolled group, p 0.229).

### Survival Analysis

A survival analysis plot (Kaplan-Meier curve) was constructed from the available diagnosis dates, death dates and follow up data. The date of diagnosis was considered the start date, and date of death the end date. There was an early and persistent divergence of the survival curves in favour of the enrolled group. Given the variable follow up periods, many patients were censored after about 700 days of follow up, limiting graph interpretability beyond that time (Figure 3).

**Figure 1:**
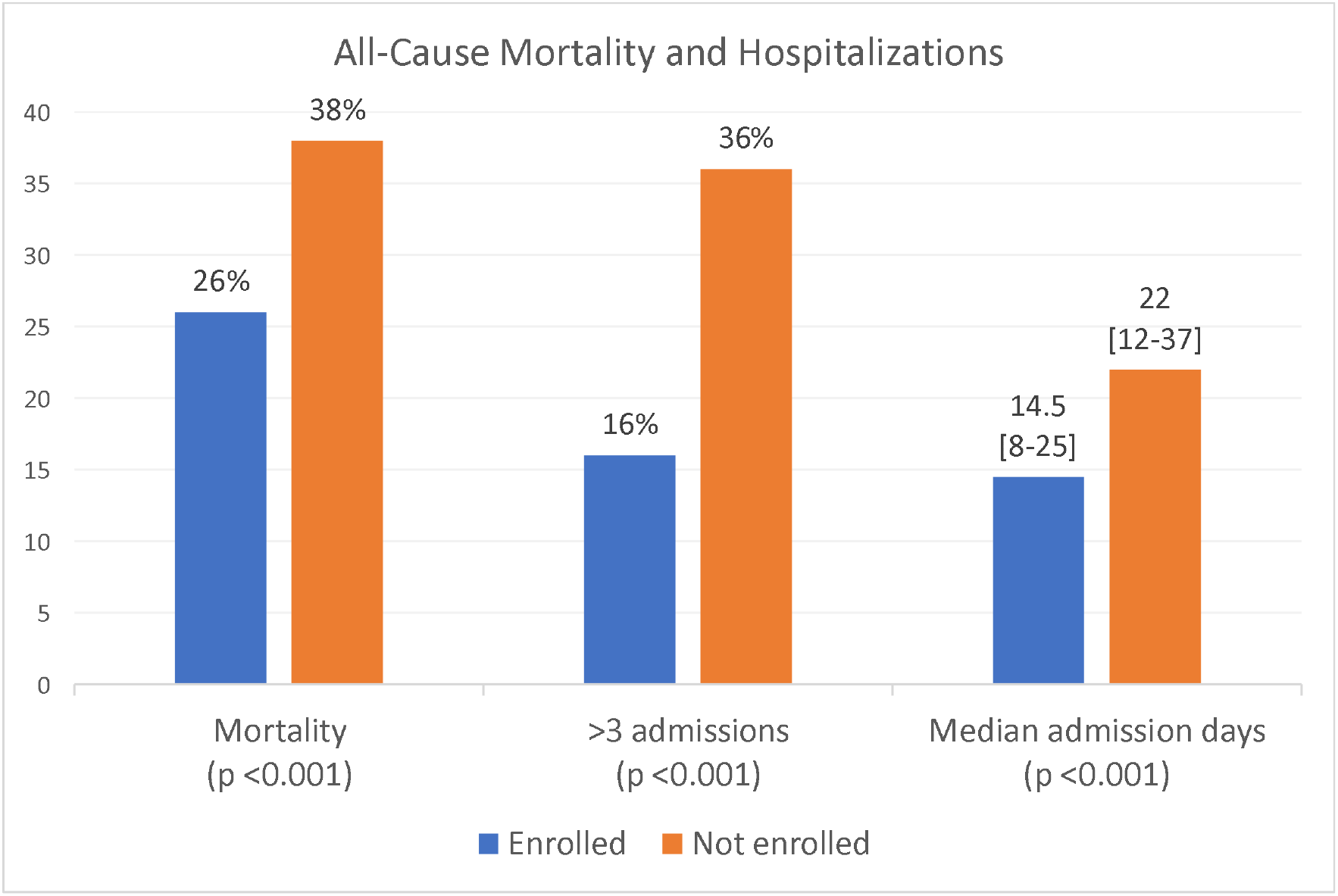
Death and hospitalizations by enrolment.

**Figure 2:**
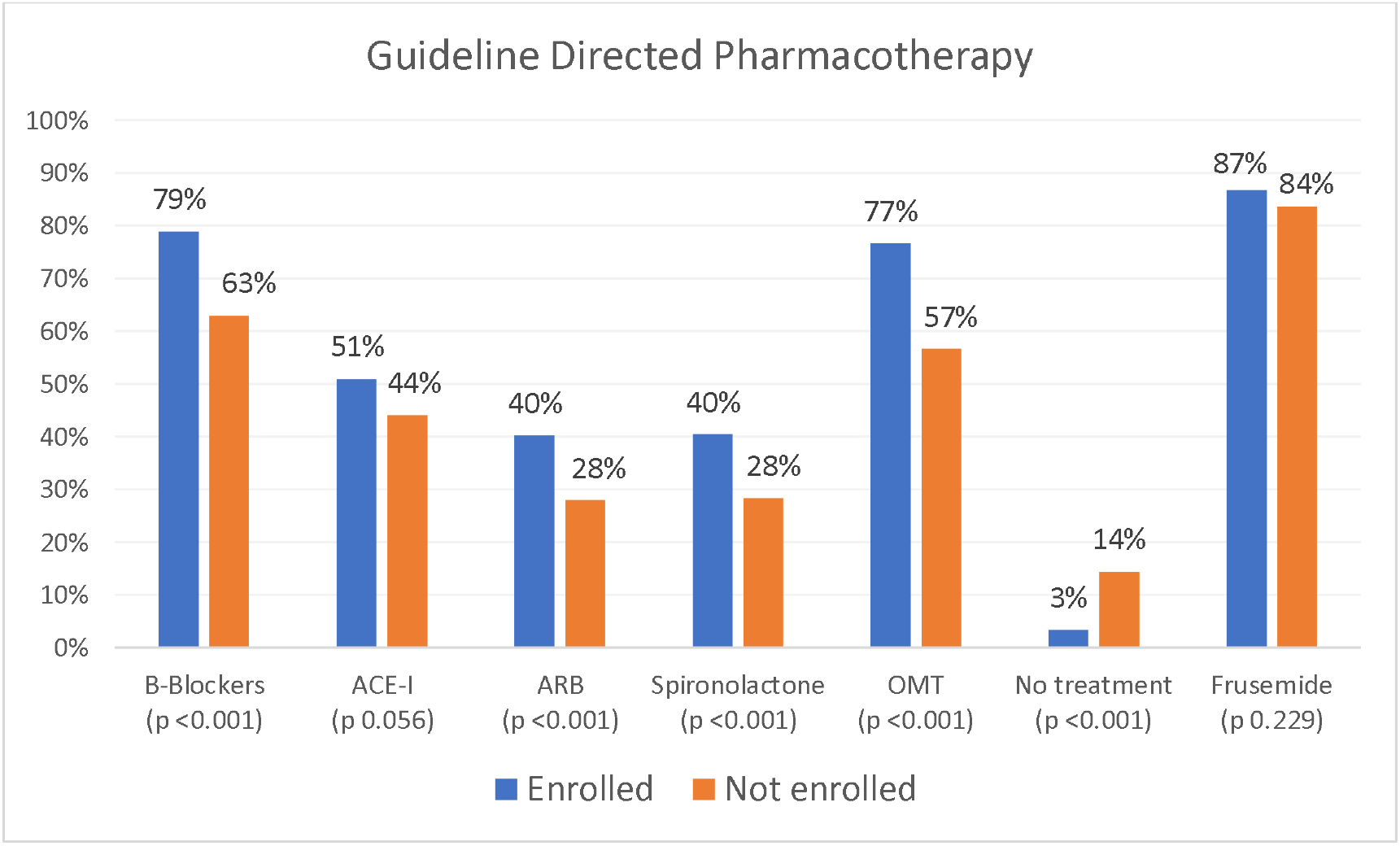
Heart failure therapy by enrolment.

**Figure 3:**
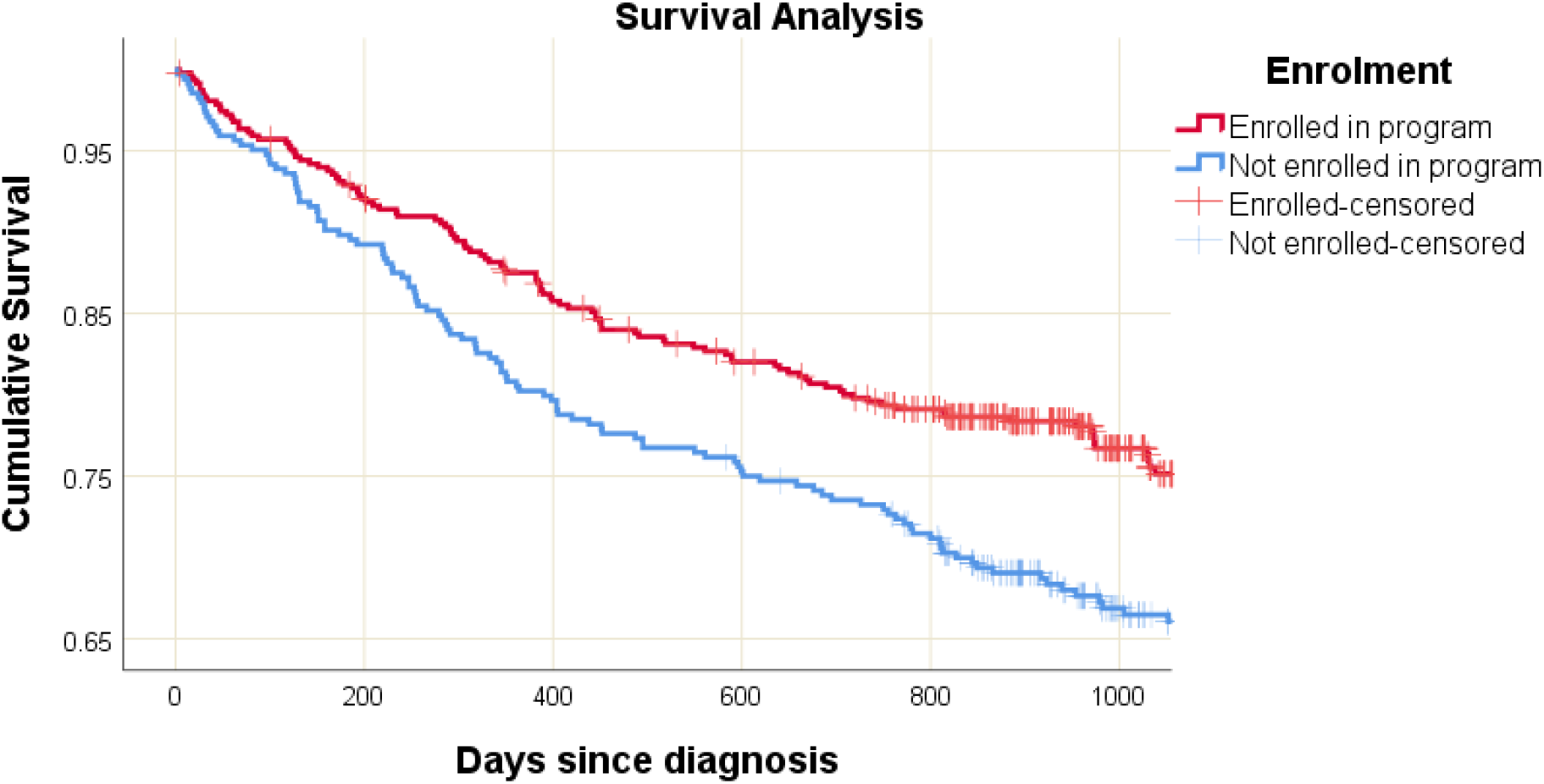
Kaplan Meier Survival Analysis.

## Discussion

Despite the relatively high mortality rate, rarely have non-pharmacological interventions been shown to improve mortality. In this this registry based study, we found that enrolment into the heart failure outreach program was significantly associated with reduced mortality, reduced frequency and length of hospital admissions as well as increased adherence to guideline directed medical therapy (GDMT). The mean age group of our study cohort is higher than much of the published literature, addressing a potential evidence gap in this group[17]. Of particular note, the enrolled cohort also had a significantly higher prevalence of ischemic cardiomyopathy (69.6% vs 49.4%, p <0.001), which generally portends a poorer prognosis than non-ischemic cardiomyopathy [18, 19]. The mortality benefit of enrolment remained statistically significant after correcting for other potential confounders such as age, medications, gender and creatinine levels.

Although previous studies have demonstrated similar reductions in hospitalisations frequency and duration, the significant mortality benefit observed in our study is in contrast to much of the published literature [20-22]. There are multiple potential explanations for this observed effect. First, our study had data over a much longer follow up period than much of the published literature. The median follow up duration in our study was 978 days. Two other studies that showed a trend for mortality benefit after longer follow up periods (of at least one year) were the ExTraMATCH meta-analysis and the HF ACTION trial [15, 16]. ExTraMATCH (Exercise training meta-analysis of trials in patients with chronic heart failure) was published in 2004, included 9 randomised trials and with a mean follow up period of 705 days. It came to the conclusion that there was clear evidence of a reduction in mortality[16]. On the other hand, ExTraMATCH II was published in 2018 and did not show such a convincing mortality benefit. Of particular interest, in ExTraMATCH II there was a more significant mortality benefit in two subgroups, older patients and those with ischemic cardiomyopathy [23]. These two patient characteristics that showed the most mortality benefit made up the overwhelming majority of our study cohort. The HF ACTION (Heart Failure and a controlled trial investigating outcomes of exercise training), which is the largest trial so far in this domain, has shown a trend for mortality benefit only after 12 months of follow up [15].

Second, the heart failure outreach service in SWSLHD is not an isolated exercise program, and to the best of our knowledge there is no available literature that studies the impact of such a personalised and integrated approach on heart failure outcomes. It is a personalised, multidisciplinary clinical nurse consultant (CNC) lead service that focuses on identifying individual patient needs, employing evidence based interventions and ensuring “getting the pills right”. This involves regular home visits patient education about the importance of medication and dietary adherence (including salt and fluid awareness and restriction), dose titration and addressing side effects and intolerances, as well as behavioural interventions, self-care, encouragement and support. All of these components are in addition to the standard supervised physical exercises tailored to the individual patient. The duration of the intervention is highly variable also, ranging from weeks to months, and in some cases even years until the patient feels confident and ready and the CNC is satisfied with the progress. Furthermore, the program can be repeated after every admission episode if needed.

There is an established reduced mortality and hospitalisations benefit to medication adherence interventions in heart failure [24, 25]. Conversely there is increased risk of hospitalisation and mortality with medication non-adherence particularly in context of contributing demographic, socioeconomic, psychological and personal factors [26-29]; SWSLHD heart failure outreach service’s focus on tackling these barriers to adherence may explain in large part the favourable results. This conclusion becomes even more plausible taking into consideration the non-enrolled group in our study were significantly under treated with optimal medications compared to the enrolled group. While there was no strong and definitive evidence that overall medication adherence was the major driver of the mortality benefit in the enrolled group, ACE Inhibitor and statin therapy association was statistically significant (p 0.049 and p <0.001, respectively). The statistically significant benefit of statin therapy was expectedly only observed in patients with underlying ischemic cardiomyopathy.

Third, in the local context, this study enrolled a diverse population from the South Western Sydney area which is considered an area of socioeconomic disadvantage, putting this vulnerable population at risk of increased adverse health outcomes [30-32].[30-32]. Outreach services such as the one studied facilitates access to essential health services, including specialist nurse review and early referral pathways. While this could provide a potential health outcome benefit, there is general lack of quality research in this area, and further research in this area is needed.

Our study has limitations. While it provided a snapshot of real world data in a socio-economically disadvantaged population, it lacked randomisation and the analysis was carried retrospectively, hence the risk of bias. The non-enrolled cohort could have declined enrolment due to a multitude of factors, including complex medical conditions, psychological and behavioural factors including motivation and depression, poor health literacy and adherence, all of which could have independently lead to the worse outcomes observed. Data was collected into the registry prospectively over a long period of time, introducing a risk of inconsistencies in data entry. The effect, if any however, is likely to be distributed across the cohorts. Furthermore, with the mortality data, as it was collected from multiple sources, including the primary health providers, there is a possibility of slight inaccuracies. However, this would lead to under-reporting of mortality in the non-enrolled cohort particularly, hence would not have changed the conclusions of the study. In addition, the personalised SWSLHD heart failure outreach service is a labour and expertise intensive program that works well within its setting, but this model of care could be difficult to replicate in other health systems with different economic structures, limiting the generalisability of our findings. Finally, growing ageing population and the rising prevalence of heart failure in the community particularly in the pace of the COVID-19 pandemic present significant challenges to such a model of care. These challenges likely could be overcome with innovative transformation leveraging ubiquitous modern technology.

## Conclusion

Enrolment in the heart failure outreach program was independently associated with reduced mortality, frequency and total length of admissions as well as adherence with optimal medical therapy. These results strongly support the implementation of personalized and multidisciplinary cardiac rehabilitation outreach services in patients with HFrEF. Given the complexity of HF and the rapidly shifting demographic, economic and technological landscape, particularly in the face of the COVID-19 pandemic, much more research is urgently needed. Incorporating insights from this study and leveraging the wealth of locally accumulated expertise with ubiquitous modern technology for enhanced communication, monitoring, education and support of this particularly vulnerable population requires urgent further research.

## Data Availability

The data used for this study is available in MS Excel/CSV format and can be shared upon request in line with institutional guidelines

